# Lithium’s effects on serum neurofilament light in Parkinson’s disease

**DOI:** 10.1101/2025.03.04.25323314

**Authors:** Thomas Guttuso, Rachel Shepherd, Daniel Sirica, Gregory E. Wilding

## Abstract

**Background:** Blood neurofilament light (NfL) is a disease-progression biomarker in Parkinson’s disease (PD).

**Objectives:** To determine the effects of lithium therapy on serum NfL in PD.

**Methods:** Frozen serum samples from 28 PD patients were assessed for serum NfL using the SIMOA platform at baseline and after 24-weeks of lithium therapy. Pairwise comparisons were performed using the Fisher-Pitman permutation test.

**Results:** Median % changes in serum NfL were -12.8, -2.0 and 11.2 in three patient groups defined by serum lithium levels at week 24: “high lithium” (0.21-0.56mmol/L, n=10), “medium lithium” (0.14-0.20mmol/L, n=8) and “low lithium” (<0.10-0.12mmol/L, n=10), respectively. Pairwise group comparisons showed significant differences between high and low lithium (*p*=0.0001) and high and medium lithium (*p*=0.0203) but not medium and low lithium groups (*p*=0.0907).

**Conclusions:** Lithium therapy achieving serum levels 0.21-0.56mmol/L significantly reduced serum NfL in PD, which strongly supports further clinical investigation of lithium’s potential disease-modifying effects in PD.

**Financial Disclosure:** Thomas Guttuso, Jr. is the President and majority owner of e3 Pharmaceuticals, Inc., which manufactures a lithium aspartate dietary supplement. None of the other authors have any financial disclosures.

**Study Funding:** This study was funded by the National Center for Advancing Translational Sciences of the National Institutes of Health under award number UL1TR001412 to the University at Buffalo.

## Introduction

Parkinson’s disease (PD) is the second most common and fastest growing neurodegenerative disorder.^1^ Although there are several FDA-approved therapies that treat the motor symptoms of PD, there remains a large unmet need for therapies that can slow the progressive neuronal degeneration and resulting progressive worsening of motor and non-motor symptoms, such as dementia, that lead to disability and loss of independence.^2-5^ One strategy to identify potential disease-modifying therapies worth advancing into later stage clinical trials is to initially focus on a therapy’s ability to positively affect biomarkers known to reflect neurodegeneration and symptom progression in PD, a.k.a. disease-progression biomarkers.

Although dopamine transporter (DaT) imaging reflects nigrostriatal tract degeneration,^6^ it does not capture degeneration in other brain sites pathologically linked to cognitive decline and dementia.^7^ Also, DaT imaging may be confounded by pharmacologic effects from dopaminergic therapy rendering it a suboptimal biomarker endpoint particularly in trials enrolling patients receiving such therapies.^8,9^ Currently, the disease-progression biomarkers shown to reflect both neuronal degeneration and progressive worsening of motor and cognitive symptoms in PD patients receiving dopaminergic therapy are free water assessed by diffusion MRI and blood neurofilament light (NfL).^10-15^ Free water reflects cell atrophy and degeneration as well as inflammation while NfL is a protein expressed exclusively in neurons and reflects any process causing neuronal damage.^16,17^

Although no therapy has been shown to reduce, i.e. improve, brain FW or blood NfL in PD, virtually all of the approved multiple sclerosis (MS) disease-modifying therapies previously assessed reduce blood NfL.^18^ Furthermore, the magnitude of NfL reduction from MS disease-modifying therapies is associated with the degree of improvement in clinical disability.^18,19^ Reduction of plasma NfL was also a critical factor resulting in FDA-approval of tofersen as a disease-modifying therapy for SOD1-amyotrophic lateral sclerosis (ALS).^20^ Thus, a therapy shown to significantly decrease brain FW in sites linked to PD progression or decrease blood NfL would represent a promising, potential disease-modifying therapy for PD worthy of further clinical investigation.

Lithium has multiple neuroprotective actions including suppressing microglial activation, reducing inflammation and oxidative stress, and enhancing autophagy and mitochondrial biogenesis and function.^21-25^ Lithium treatment has demonstrated benefit in several PD animal models.^26-29^ Also, the 77% risk reduction of PD in smokers shown in prospective cohort studies has been theorized to be due to the high levels of lithium in tobacco.^30^ Based on this background, our group performed a pilot clinical trial in PD.^31^ Results showed 24 weeks of lithium aspartate therapy 45mg/day to be associated with the largest and most consistent reductions in FW in three brain sites where FW progression is known to reflect worsening motor (posterior substantia nigra (pSN))^10,32^ and cognitive symptoms (nucleus basalis of Meynert (nbM) and dorsomotor nucleus of the thalamus (DMN-T)).^11,33^ The same lithium dosage also maximally engaged the PD therapeutic target nuclear receptor-related 1 protein (Nurr1).^34-36^ Although these results were encouraging, our pilot study provided 24-week data on 17 PD patients of whom only 4 received the most promising lithium dosage of 45mg/day. As a result, another trial was needed to further explore lithium’s effects on these biomarkers as well as blood NfL.

Here we report on the serum NfL results from both this study and frozen samples from the previous pilot study^31^ while awaiting other blood-based and neuroimaging biomarker results including brain FW. Serum glial fibrillary acid protein (GFAP) was also assessed as it was included by the vendor for no additional cost and may reflect and predict cognitive impairment in PD.^37^

## Methods

This trial (ClinicalTrials.gov ID: NCT06099886) enrolled 15 patients, aged 45-80, diagnosed with PD by UK Brain Bank Criteria for <4 years^38^. PD or psychiatric medication dosages needed to be stable for >30 or >60 days, respectively; patients could have no history of brain surgery, stroke, use of lithium or antipsychotic medication; needed a Montreal Cognitive Assessment (MoCA) score >20, normal thyroid stimulating hormone (TSH) level and estimated glomerular filtration rate ≥50 at screening. Our previous pilot study had the same patient eligibility criteria with the exception that PD patients of any disease duration were eligible.^31^

All 15 patients provided written informed consent. At the baseline (BL) visit, patients fasted for 10-12 hours before providing a venous blood sample. Serum was stored at -70C until NfL and GFAP were assessed in duplicate by *Quanterix* (Lexington, MA) using the SIMOA platform. Safety laboratory tests included a complete blood count, comprehensive metabolic panel including serum calcium and thyroid stimulation hormone levels (*Kaleida Health Laboratories*, Williamsville, NY). After obtaining an MRI scan of the head, all patients were placed on lithium aspartate 5mg capsules starting at 10mg, 2x/day with the dosage increased by 10mg/day every week up to 45mg/day (20mg qAM and 25mg qhs) for the remainder of the 24-week treatment period. This lithium titration was used to maximize tolerability. At the 24- week visit, another fasting blood sample was obtained and processed the same as for the BL visit and serum lithium was assessed 10-12 hours after the last lithium dose.

After reviewing the serum NfL results, the 15 patients from the current study and 13 patients from the previous pilot study for whom serum was available were divided into three groups based on serum lithium levels at the 24-week visit: “high lithium” (0.21-0.56mmol/L, median=0.32mmol/L, *n*=10), “medium lithium” (0.14-0.20mmol/L, median=0.17mmol/L, *n*=8) and “low lithium” (<0.10-0.12mmol/L, median<0.10mmol/L, *n*=10). Pairwise comparisons were performed using the Fisher-Pitman permutation test.

Data Sharing: All data produced in the present study are available upon reasonable request to the authors.

## Results

Sixteen patients were enrolled from October 2023-June 2024. One patient withdrew prior to starting lithium therapy. Four patients receiving lithium reported mild adverse events of sedation, nausea or dyskinesia that did not require lithium dosage reduction. There were no clinically meaningful changes in any safety laboratory values.

Patient BL characteristics are summarized in Table 1. Patients in the low lithium group were slightly younger, had fewer females, longer disease duration and higher levodopa equivalent daily dose.

**Table 1:** Patient Baseline Characteristics

|  | High Lithium (*n*=10) | Medium Lithium (*n*=8) | Low Lithium (*n*=10) |
| --- | --- | --- | --- |
| Age (years) | 65.5 (8.4) | 68 (5.5) | 62 (6.6) |
| % Female | 60 | 62.5 | 30 |
| Disease Duration (years) | 1.75 (1.79) | 1.75 (1.24) | 4.5 (4.29) |
| MDS-UPDRS | 21.5 (12.9) | 18 (12.8) | 21.5 (14.8) |
| MoCA | 26 (3.1) | 28.5 (2.0) | 29 (2.6) |
| LEDD (mg) | 425 (236) | 425 (280) | 750 (577) |
| Serum NfL (pg/ml) | 16.45 (7.83) | 14.2 (3.44) | 16.6 (7.08) |
| Serum GFAP (pg/ml) | 204 (191) | 134 (41) | 153 (125) |

Values are median (standard deviation), MDS-UPDRS: Movement Disorder Society-Unified Parkinson’s Disease Rating Scale, MoCA: Montreal Cognitive Assessment, LEDD: Levodopa Equivalent Daily Dose, NfL: Neurofilament Light, GFAP: Glial Fibrillary Acidic Protein.

**Figure 1:**
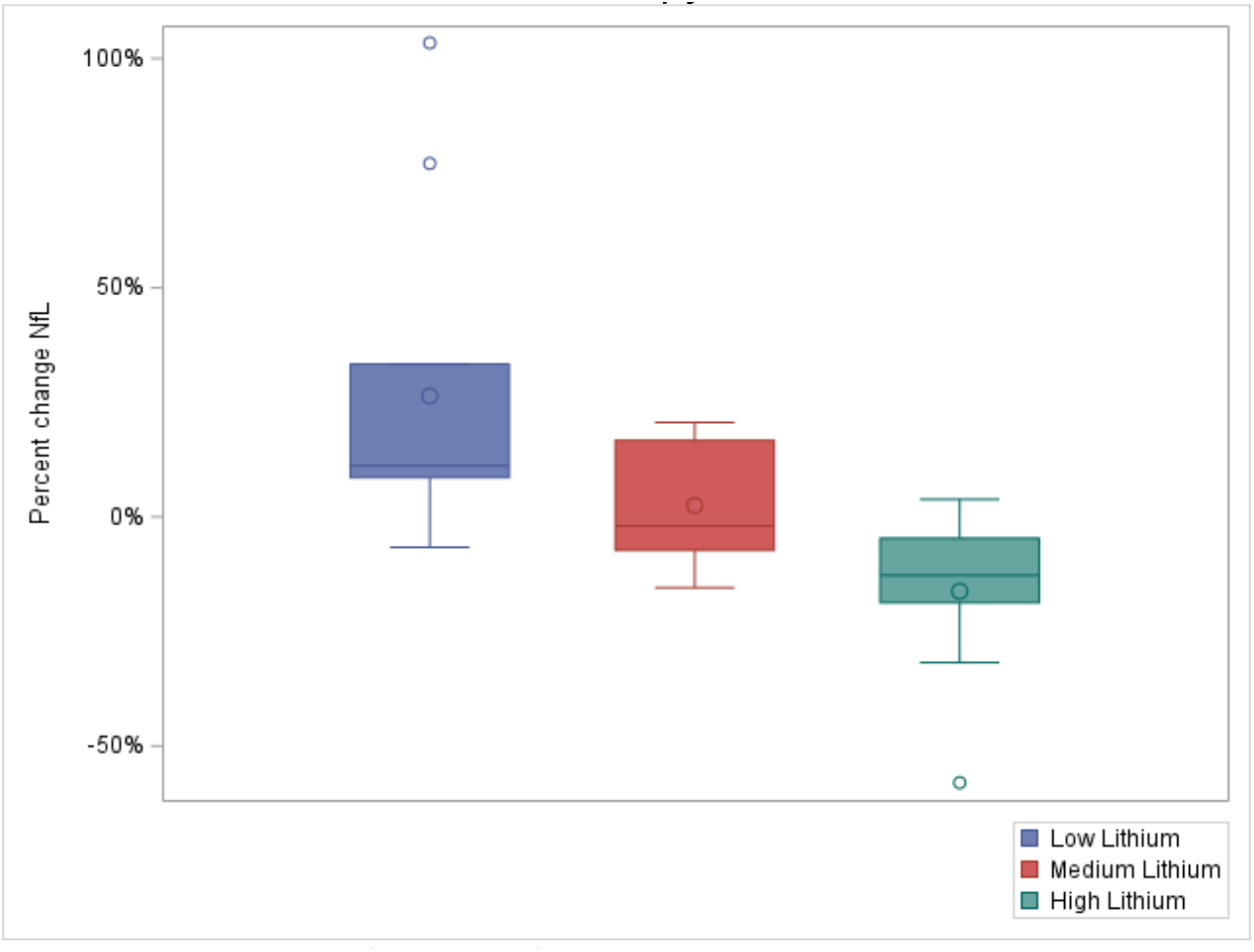
% Change in Serum NfL after 24-Weeks of Lithium Therapy. Each colored box reflects the first to third interquartile range. Lines and dots in each box are median and mean values, respectively. Dots outside the boxes are patient outliers. Whiskers mark the smallest and largest data points within 1.5x of the first and third interquartile range, respectively. Intergroup comparisons: high versus low lithium (*p*=0.0001), high versus medium lithium (*p*=0.0203), medium versus low lithium (*p*=0.0907).

Median % changes in serum GFAP were 7.3, 42.8 and 12.4 in the high, medium and low lithium groups, respectively. Pairwise comparisons showed a significant difference between the high and medium lithium groups (*p*=0.0075) but not the high and low lithium groups (*p*=0.1763) or the medium and low lithium groups (*p*=0.3950).

## Discussion

To our knowledge, this is the first report of lithium therapy leading to a significant decrease in serum NfL in any patient population and the first report of any therapy reducing serum NfL in PD. Although the results are only based on 28 patients, they do show a clear dose-response relationship between serum lithium levels and reductions in serum NfL (Figure 1). The pairwise comparisons indicate the high lithium group, with serum lithium levels of 0.21- 0.56mmol/L, as leading to the largest reductions in serum NfL. Although the low lithium group had some baseline differences compared to the other two groups, there still was a significant difference between the high and medium lithium groups, which had very similar baseline characteristics (Table 1).

It is unclear if the 12.8% median reduction in serum NfL seen in the high lithium group would be sufficient to reflect slowing of clinical progression and disability in PD; however, comparisons with MS and ALS therapies may provide insight. Although MS has a much different pathophysiology than PD, primary progressive MS (PPMS) has a temporal profile more similar to PD than relapsing-remitting MS. Ocrelizumab is currently the only approved disease-modifying therapy for PPMS and reduces plasma NfL by 15% after 24-weeks.^19^ In ALS, an experimental therapy has recently been shown to significantly reduce long-term, all-cause mortality by 60% and plasma NfL by 10% relative to placebo after 24-weeks.^39^ Based on these comparisons, the 12.8% reduction in serum NfL from lithium therapy reported here is of a magnitude that could potentially reflect disease-modifying benefit in PD.

Although there was significantly less of an increase in serum GFAP in the high lithium group compared to the medium group, there was not a clear dose-response relationship across the 3 lithium groups nor a significant difference between the high and low lithium groups, as was seen for changes in serum NfL (Figure 1).

Further clinical research needs to be performed to determine if these serum NfL findings are consistent in a larger number of PD patients receiving lithium, the durability of these effects over longer durations of treatment and the effects of lithium dosages achieving higher serum lithium levels on serum NfL both at the group and individual patient levels. One of us previously reported anecdotal reductions in off-time in PD patients placed on adjunct lithium therapy with maximum subjective reductions reported with serum lithium levels of 0.4-0.5mmol/L and reduced tolerability with levels >0.60mmol/L.^40^ Based on these experiences, we recently initiated an extension study where patients completing the first 24-week lithium trial were eligible to receive an additional 24-weeks of lithium therapy with the dosage titrated to achieve a target serum lithium level of 0.4-0.5mmol/L (Clinicaltrials.gov ID: NCT06592014). Results from this extension study will provide preliminary data to help answer the above questions. We also initiated a small randomized controlled trial (RCT) that will enroll 20 additional PD patients, 10 of whom will receive placebo and 10 lithium (Clinicaltrials.gov ID: NCT06339034); all 20 patients will be eligible to enroll in the extension study.

The most important question regarding lithium’s ability to provide disease-modifying benefit for PD can only be answered through a much larger and longer duration RCT assessing both biomarker and key clinical outcomes such as cognitive impairment and postural instability that are highly disabling in PD and usually are unresponsive to dopaminergic therapy.

## Conclusion

In this pooled analysis from two small clinical trials, lithium therapy achieving serum levels of 0.21-0.56mmol/L was associated with a significant 12.8% reduction in serum NfL in PD. Considering that blood NfL is an established disease-progression biomarker in PD and a validated disease-modifying therapeutic biomarker in MS and potentially ALS, further research is merited investigating lithium’s potential disease-modifying effects in PD.

## Data Availability

All data produced in the present study are available upon reasonable request to the authors.

